# Socioeconomic profile of people affected by skin neglected tropical diseases in the communes of Zagnanado and Allada, Benin: a mixed-methods cross-sectional study

**DOI:** 10.64898/2026.04.28.26351648

**Authors:** Roch Houngnihin, Mélissa Biao, Bernice Gbèbioho, Fréjus Togonou, Daniel Azonchiga

## Abstract

**Background:** Skin neglected tropical diseases (skin NTDs) including Buruli ulcer, leprosy, lymphatic filariasis, and scabies disproportionately affect impoverished rural communities in sub-Saharan Africa. In Benin, their persistence despite two decades of national control programmes highlights the need for locally grounded socioeconomic evidence. We characterised the socioeconomic profile of people affected by skin NTDs in two endemic communes and examined commune-level disparities in access to care and the socio-ecological factors driving transmission.

**Methodology/Principal Findings:** We conducted an explanatory sequential mixed-methods cross-sectional study in the communes of Zagnanado (Zou department) and Allada (Atlantique department), Benin, between November 2024 and May 2025, following STROBE guidelines. The quantitative component enrolled 403 participants (rights-holders and their relatives) through weighted proportional random sampling from the catchment areas of two Buruli ulcer screening and treatment centres (CDTUBs), with an effective participation rate of 99.8%. The qualitative component comprised 60 purposively selected semi-structured interviews. The sample was balanced by sex (50.6% women; 49.4% men) and dominated by adults aged 25–49 years (55.1%). Participants were predominantly engaged in farming, livestock-rearing, or fishing (43.7%), lived in rural areas (50.1%), and had low educational attainment (38.5% with no formal schooling). Treatment cost was the leading barrier to care (84.6%), with no significant commune difference (χ²=0.62, p=0.43). By contrast, limited geographical access (Allada 26.4% vs. Zagnanado 66.7%; χ²=65.7, p<0.001) and inadequate health infrastructure (10.0% vs. 53.0%; χ²=88.8, p<0.001) showed marked intercommunal disparities. Critically, Buruli ulcer was spontaneously recognised by only 7.2% of respondents despite both study sites hosting specialised treatment centres a finding we term “nosological dissociation”. Qualitative data revealed widespread mystical illness interpretations (22.8% attributed skin disease to witchcraft or curses) and a plurality of Fongbé vernacular terms that perceptually disconnect biomedical conditions from their local names.

**Conclusions/Significance:** Skin NTDs in these two Beninese communes affect impoverished rural populations whose informal livelihoods expose them to hydromorphic environments. Financial and infrastructural barriers operate differently across communes, warranting context-specific responses: financial protection mechanisms in Allada, service availability strengthening in Zagnanado, and improved water and sanitation in both. Structured collaboration with traditional medicine practitioners strongly endorsed by 78.9% of participants and culturally adapted awareness campaigns using vernacular disease terminology are essential to close the nosological dissociation gap and reduce delays in care-seeking.

**AUTHOR SUMMARY:** Skin neglected tropical diseases such as Buruli ulcer and leprosy cause severe disability and persistent poverty in Benin’s rural communities. Despite specialised treatment centres operating in two endemic communes (Zagnanado and Allada), Buruli ulcer was spontaneously named by only 7% of the 403 people we surveyed and many patients knew only its local Fongbé names such as *akpa djɔmakou* or *timantibo*, without connecting it to its biomedical identity. We also found that most of those affected are subsistence farmers and fishers working in waterlogged environments, unable to pay for biomedical treatment (84.6%) and often turning first to traditional healers because they interpret skin wounds as curses or divine punishment. Importantly, the barriers to care differ between the two communes: the more urban Allada needs financial support programmes, while the more remote Zagnanado needs investment in health infrastructure. Our findings argue for a three-pronged response: financial protection for patients; awareness campaigns that bridge vernacular and biomedical disease concepts; and a formal partnership between health systems and traditional healers the first port of call for most affected families.

## INTRODUCTION

Population health is fundamentally shaped by social, economic, and environmental conditions. The World Health Organization (WHO) estimates that social determinants of health including education, income, housing, and access to water and sanitation account for 30% to 55% of differences in health outcomes globally [1]. In 2022, approximately 719 million people (9.2% of the global population) lived on less than US$2.15 per day [2], directly constraining their capacity to access care, adequate nutrition, and decent living conditions.

Among the diseases flourishing in these contexts of deprivation are the neglected tropical diseases (NTDs), a heterogeneous group of infections and infestations that proliferate in poor tropical communities [3]. Skin NTDs including Buruli ulcer (*Mycobacterium ulcerans* infection), leprosy, cutaneous leishmaniasis, onchocerciasis, lymphatic filariasis, scabies, yaws, and mycetoma [4] cause not only physical suffering but also lasting disability, profound social stigma, and loss of livelihood. Each year, an estimated 1.65 billion people require preventive or curative NTD interventions [5].

In Benin, skin NTDs persist despite the sustained efforts of the National Programme for the Control of Leprosy and Buruli Ulcer (PNLLUB) and the PI-PSILUB I and II projects [6]. These diseases disproportionately affect rural areas characterised by structural poverty, where populations rely mainly on farming, livestock-rearing, and fishing in environments conducive to pathogen transmission. The communes of Zagnanado (Zou department) and Allada (Atlantique department) are endemic zones where the presence of specialised Buruli ulcer screening and treatment centres (CDTUBs) attests to the burden of these conditions. Limited access to health infrastructure, entrenched traditional beliefs, and frequent recourse to traditional medicine hinder awareness campaigns, early detection, and adequate clinical management [7].

This study is part of the DESABEN-MTN project (Social Determinants of Health and Neglected Tropical Diseases), implemented through the Fondation Anesvad, the Fondation Raoul Follereau, and the Laboratoire d’Anthropologie Médicale Appliquée (LAMA) of the University of Abomey-Calavi. It responds to a documented gap: while commune-level epidemiological data on skin NTDs in Benin are scarce, designing effective and equitable control programmes requires detailed understanding of the socioeconomic characteristics, living conditions, and care-seeking behaviours of affected populations.

### Objectives

The general objective was to document the socioeconomic profile of people affected by skin NTDs in the communes of Zagnanado and Allada. Specific objectives were to: (i) describe sociodemographic characteristics (age, sex, education, occupation, ethnicity); (ii) identify main income sources and characterise living conditions (housing, water, sanitation); (iii) examine commune-level disparities in access to care; and (iv) explore the sociocultural representations and environmental factors shaping transmission and health-seeking behaviour.

## MATERIALS AND METHODS

### Study setting

The study was conducted in two rural communes in southern and central Benin (Fig 1). **Zagnanado** (Zou department) covers 750 km² and has approximately 55,061 inhabitants [8]. Its population is predominantly Mahi and Yoruba, with a mix of Christian and traditional religions. The commune has one CDTUB and six *arrondissements*. **Allada** (Atlantique department) covers 381 km² and has 125,712 inhabitants [8]. The Aïzo (83%) and Fon (10%) ethnic groups predominate, with a strong presence of traditional religion (63.9%). The commune is subdivided into 12 *arrondissements* and hosts a departmental CDTUB specialised in the management of Buruli ulcer, leprosy, and other skin NTDs. The two communes were selected because both host operational CDTUBs, they represent distinct socioeconomic and geographic profiles (peri-urban vs. remote rural), and both were identified as priority intervention zones by the DESABEN-MTN project.

**Fig 1.**
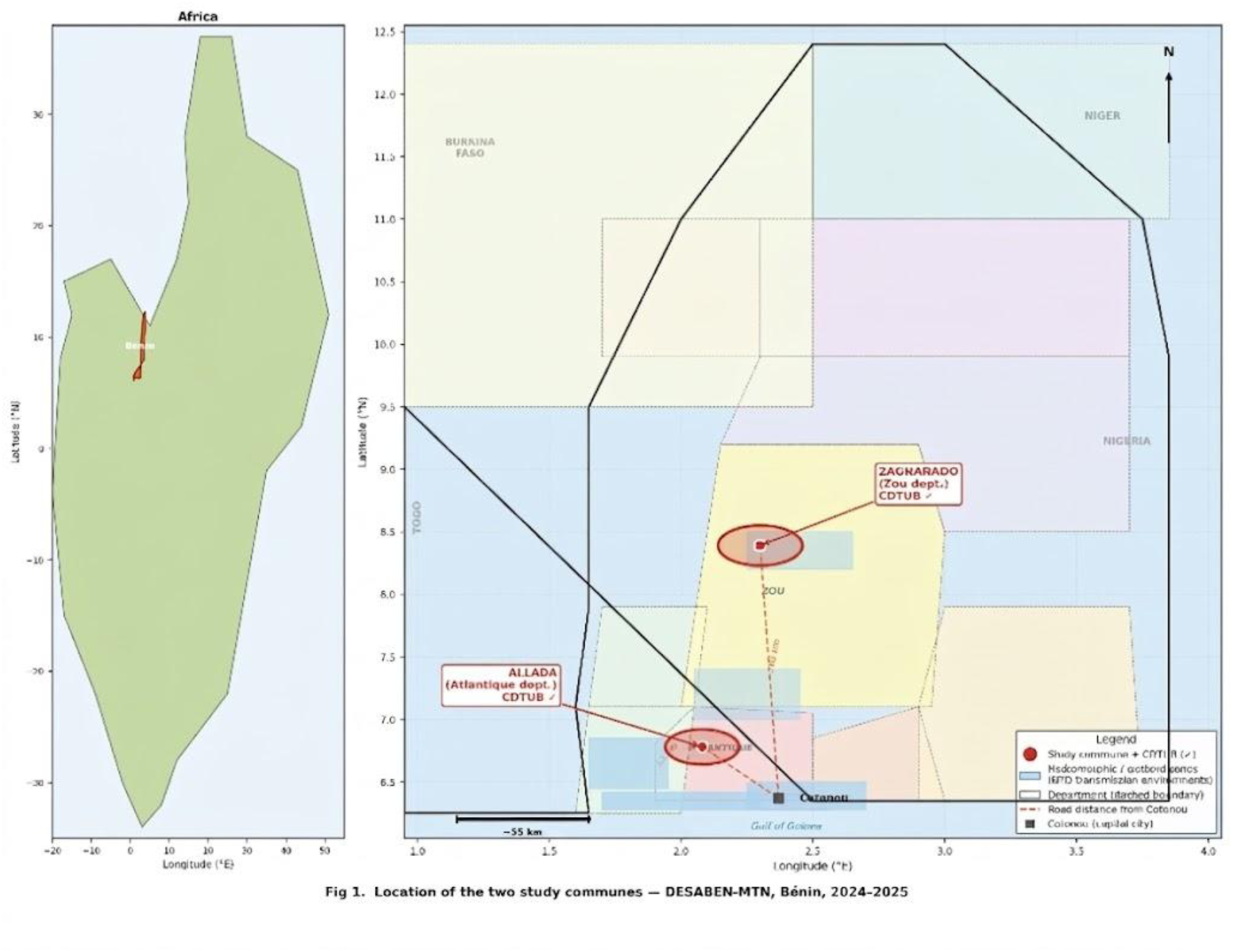
Location of the two study communes, DESABEN-MTN, Bénin, 2024–2025. Red circles = study communes (Allada, Atlantique department; Zagnanado, Zou department), both hosting Buruli ulcer screening and treatment centres (CDTUB). Dark blue areas = Lake Nokoué, coastal lagoon, and Lake Ahémé. Light blue areas = hydromorphic / wetland zones (key NTD transmission environments). Dashed orange lines = approximate road distances from Cotonou. Dashed grey lines = department boundaries. Data sources: GADM; INSAE RGPH4 (2013); OpenStreetMap contributors.

### Conceptual framework

The study drew on the WHO conceptual framework of social determinants of health [9], adapted to the context of skin NTDs. Three dimensions were considered: (i) sociocultural determinants (beliefs, norms, traditional practices, education); (ii) socioeconomic determinants (poverty, access to care, employment, income sources); and (iii) environmental factors (access to drinking water, hygiene, sanitation, housing, climate). The One Health approach, which recognises the interdependence of human, animal, and environmental health, further guided the analysis [10].

### Study design

DESABEN-MTN is an explanatory sequential mixed-methods cross-sectional study [11], in which quantitative data were collected and analysed first and then complemented by the qualitative component.

This design was chosen because the objectives required both a statistical profile of the affected population and an exploration of the social logics, representations, and practices underlying observed health behaviours. The cross-sectional observational study was conducted in accordance with the STROBE reporting guidelines (S1 Checklist).

### Population, sampling, and participant classification

Three categories of actors were identified: (i) rights-holders persons with clinical signs of skin NTDs and their household relatives, whether or not they attended CDTUBs; (ii) duty-bearers formal health providers and traditional healers; and (iii) obligation-bearers local authorities, PNLLUB officials, and departmental health directorates.

Among the 403 quantitative participants, participants were recruited both from CDTUB patient registers (confirmed or suspected skin NTD cases) and from household or community members through a door-to-door census in surrounding villages. This distinction is critical for interpretation, as the inclusion of both patients and their relatives was intentional, allowing programme planners to understand not only individuals already engaged in care but also those who do not access health services.

Sample size was calculated using the Schwartz (1995) formula, yielding 404 participants after a 5% safety margin. Proportional random sampling was applied to residents of CDTUB catchment areas, with allocation proportional to the number of *arrondissements* per commune: 12 in Allada (220 participants, 54.6%) and 6 in Zagnanado (183 participants, 45.4%). The effective participation rate was 99.8% (403/404), with a single refusal before the interview.

Inclusion criteria: being affected by a skin NTD or being a household relative of an affected person; residing in the catchment areas of the CDTUBs of Zagnanado or Allada. Non-inclusion criteria: absence of personal or household NTD exposure; being affected by unrelated conditions only.

For the qualitative component, non-probability purposive sampling was used to select 60 participants: 36 rights-holders, 14 duty-bearers, and 10 obligation-bearers. Saturation was assessed iteratively and reached after approximately 45 interviews, with the final 15 conducted to confirm thematic stability.

### Data collection

Quantitative data were collected using a structured questionnaire administered on Android smartphones via ODK Collect version v2023.2 (Open Data Kit; https://getodk.org/). Variables included sociodemographic characteristics, economic activities, income sources, NTD knowledge, and access to care indicators. The questionnaire was pre-tested with 20 participants in a non-study commune (Abomey-Calavi) to assess item comprehension, administration duration, and internal consistency; ambiguous items were reformulated before definitive deployment.

Qualitative data were collected through semi-structured interviews conducted in Fongbé (the predominant local language) and French, audio-recorded with participant consent, and transcribed verbatim. A structured direct observation grid documented living conditions, hygiene practices, and the physical environment of participants.

### Data analysis

Quantitative data were analysed using IBM SPSS Statistics version 29.0 (IBM Corp., Armonk, NY, USA). Descriptive statistics (frequencies, percentages, means) characterised the sample. Pearson chi-squared tests examined commune-level associations for access barriers (Table 3) and proposed control strategies (Table 6). Where expected cell counts fell below 5, Fisher’s exact test would have been applied; no such cases were observed. Cramér’s V was calculated to assess effect size (V<0.10 negligible, 0.10–0.30 small, 0.30–0.50 moderate, >0.50 large). A significance threshold of α=0.05 was applied throughout. Crude odds ratios (OR) with 95% confidence intervals (95% CI) were computed from 2×2 contingency tables using Woolf’s method for key bivariate associations (Table 7).

Qualitative data were analysed using Braun and Clarke’s (2006) reflexive thematic analysis, conducted manually. The six-phase process comprised: (1) data familiarisation through repeated reading of transcripts; (2) initial code generation; (3) theme searching; (4) theme review; (5) theme definition and naming; and (6) report production. Triangulation across quantitative results, interview data, and direct observation enabled convergent interpretation and cross-validation of findings. Two researchers independently coded 20% of the corpus; inter-rater agreement was assessed and discrepancies resolved through discussion.

### Ethics

This study was conducted in accordance with the principles of the Declaration of Helsinki (World Medical Association, 2013 revision). Ethical approval was granted by the National Ethics Committee for Health Research of Benin (Comité National d’Éthique pour la Recherche en Santé, CNERS), under reference CNERS 002/2025. Free and informed written consent was obtained from all adult participants prior to enrolment. For participants under 18 years of age, written informed consent was obtained from a parent or legal guardian; assent was additionally sought from adolescents aged 12 years and over. Anonymity and data confidentiality were ensured throughout by assigning unique pseudonyms to each participant. Consent to publish direct quotations in anonymised form was obtained at the time of interview.

## RESULTS

### Sociodemographic characteristics

The 403 participants were almost evenly distributed between women (50.6%) and men (49.4%). The majority (55.1%) were adults aged 25–49 years, followed by participants under 25 (27.5%) and those aged 50 or over (17.3%). Table 1 summarises sociodemographic characteristics.

**Table 1.** Sociodemographic characteristics of participants (N=403). All participants enrolled between November 2024 and May 2025 in Zagnanado and Allada, Benin.

| Variable / Category | n | % |
| --- | --- | --- |
| Sex |  |  |
| Male | 199 | 49.4 |
| Female | 204 | 50.6 |
| Age group |  |  |
| Youth (< 25 years) | 111 | 27.5 |
| Adults (25–49 years) | 222 | 55.1 |
| Seniors (≥ 50 years) | 70 | 17.3 |
| Religion |  |  |
| Christian | 249 | 61.8 |
| Traditional / Endogenous | 119 | 29.5 |
| Muslim | 6 | 1.5 |
| Not stated | 29 | 7.2 |
| Ethnicity |  |  |
| Fon | 312 | 77.4 |
| Holi | 51 | 12.7 |
| Goun | 30 | 7.4 |
| Other | 10 | 2.5 |
| Place of residence |  |  |
| Rural | 202 | 50.1 |
| Semi-urban | 159 | 39.5 |
| Urban | 42 | 10.4 |
| Education level |  |  |
| None | 155 | 38.5 |
| Primary | 111 | 27.5 |
| Secondary | 120 | 29.8 |
| University | 17 | 4.2 |

Source: DESABEN-MTN survey, May 2025.

Christianity predominated (61.8%), followed by traditional/endogenous religions (29.5%) and Islam (1.5%). A marked religious syncretism where traditional practices coexist with Christianity or Islam shapes therapeutic itineraries and frequently orients patients towards traditional medicine before biomedical care. The Fon ethnic group dominated the sample (77.4%), which partly explains the plurality of Fongbé vernacular NTD terms recorded. Half of participants (50.1%) lived in rural areas, 39.5% in semi-urban settings, and only 10.4% in urban centres. Educational attainment was low: 38.5% had received no formal education and only 4.2% had reached university level.

### Economic activities and income sources

Farming, livestock-rearing, and fishing activities predominated (43.7%), followed by trade (29.0%) and civil-service employment (12.4%). A further 14.9% were unemployed or engaged in marginal activities (Table 2). The occupational distribution did not differ significantly between the two communes (chi-squared test: p=0.29).

**Table 2.** Occupational distribution of participants, by commune.

| Occupation | Allada n (%) | Zagnanado n (%) | Total n (%) |
| --- | --- | --- | --- |
| Farming, livestock, fishing | 89 (40.5) | 87 (47.5) | 176 (43.7) |
| Trade | 68 (30.9) | 49 (26.8) | 117 (29.0) |
| Civil servant | 32 (14.5) | 18 (9.8) | 50 (12.4) |
| Unemployed / other | 31 (14.1) | 29 (15.9) | 60 (14.9) |
| Total | 220 (100) | 183 (100) | 403 (100) |

Source: DESABEN-MTN survey, May 2025.

Qualitative interviews revealed that income sources are predominantly informal: subsistence agriculture, fishing, market gardening (vegetables, rice), handicrafts, and petty trade. In Allada, the collection of raffia in swamps for mat-making is a gender-specific activity carried out mainly by women that brings them into sustained contact with hydromorphic environments. Non-fixed and irregular incomes in the informal sector severely constrain households’ capacity to pay for biomedical care. Daily activities working in lowlands, streams, and swamps expose participants to reservoirs of skin NTD pathogens, and wounds caused by agricultural tools (hoes, machetes) constitute common portals of entry for infection.

### Barriers to health-care access and intercommunal disparities

Economic factors were the leading barriers to care. Treatment cost was the most frequently reported barrier overall (84.6%), without significant difference between communes (χ²=0.62, p=0.43, V=0.04). This indicates that financial unaffordability is a universal burden in both settings. By contrast, significant and large intercommunal disparities were observed for limited geographical access (Allada 26.4% vs. Zagnanado 66.7%; χ²=65.7, p<0.001, V=0.40) and inadequate health infrastructure (10.0% vs. 53.0%; χ²=88.8, p<0.001, V=0.47) (Table 3). Crude odds ratio analyses (Table 7) confirm that residents of Zagnanado were 5.59 times more likely to report limited geographical access (OR 5.59, 95% CI 3.63–8.58) and 10.15 times more likely to report inadequate infrastructure (OR 10.15, 95% CI 5.98–17.21) compared to Allada residents. Overall, only 40.7% of participants had ever used formal health services for skin NTDs.

**Table 3.**
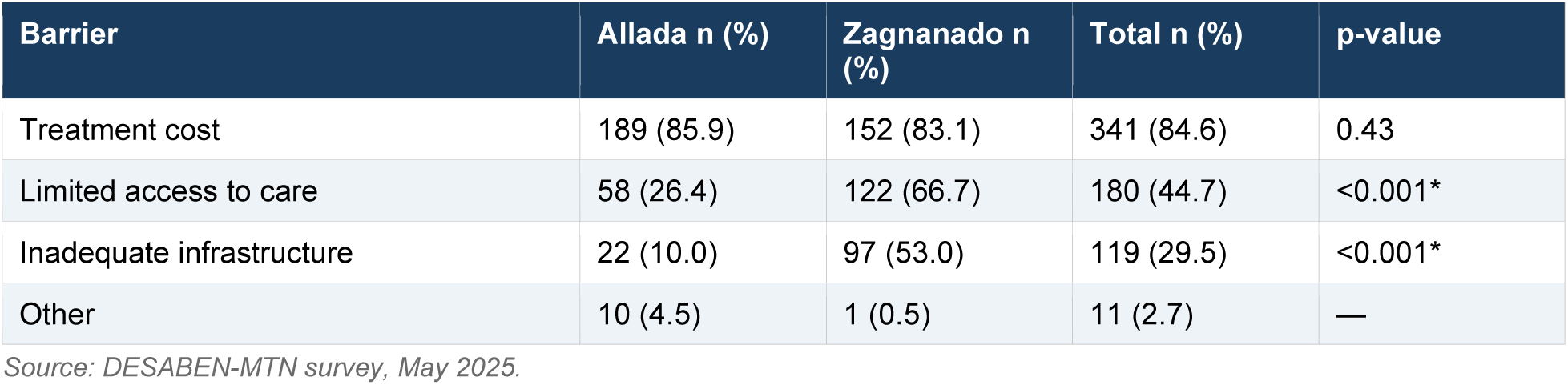
Economic and structural barriers to health-care access, by commune. p-values from Pearson chi-squared tests (2×2 contingency, barrier yes vs. no). *Significant at p<0.001. V = Cramér’s V effect size.

Qualitative testimonies illuminate these structural differences. A patient from Allada (41-45 years) explained: *“I cannot walk all the way to Allada where they treat wounds. Even with a motorbike, you need fuel and someone to take you. When you cannot even eat, how can you go there without money?”* A patient from Zagnanado (31-35 years) stated: *“The problem is that if your family has no money you cannot dare think of going to hospital. There, they will ask for money first, and without money you can do nothing. So you can only look for what is affordable — a healer who can start treatment for 2,000 FCFA and wait for the rest.”* These accounts illustrate how financial and geographical barriers intersect with differential service availability, producing distinct care-access profiles in each commune.

### A critical gap: Buruli ulcer recognition and nosological dissociation

A striking finding was the extremely low spontaneous recognition of Buruli ulcer: only 7.2% of participants named it, despite both study sites hosting CDTUBs specifically dedicated to its management (Table 4). NTDs most frequently cited were those targeted by mass drug administration campaigns: lymphatic filariasis (53.3%), onchocerciasis (43.4%), and sleeping sickness (41.2%). Leprosy was named by only 19.4% of respondents.

**Table 4.** Neglected tropical diseases spontaneously mentioned by participants (multiple responses allowed, N=403).

| NTD spontaneously mentioned | n | % |
| --- | --- | --- |
| Lymphatic filariasis | 215 | 53.3 |
| Onchocerciasis | 175 | 43.4 |
| Sleeping sickness | 166 | 41.2 |
| Leishmaniasis | 112 | 27.8 |
| Leprosy | 78 | 19.4 |
| Chagas disease | 76 | 18.9 |
| Buruli ulcer | 29 | 7.2 |
| Other (ringworm, chickenpox, measles, etc.) | 80 | 19.9 |

Source: DESABEN-MTN survey, May 2025.

We interpret this recognition gap as a “nosological dissociation”: the perceptual separation between a biomedical disease entity and its vernacular equivalents. Participants recognised *akpa djɔmakou* (non-healing wound), *timantibo* (tomato gri-gri), or *noudido* (cast spell) as distinct conditions from “Buruli ulcer”, despite the fact that they describe the same pathology at different stages. Several CDTUB inpatients reported never having heard of Buruli ulcer before admission, despite prior awareness campaigns in their villages. For leprosy, the local term *azon vo* (red-spot disease) was similarly disconnected from its biomedical name. These data suggest that existing awareness campaigns, centred on biomedical nomenclature, are failing to connect with local illness realities.

### Knowledge, representations, and perceived causes of skin NTDs

Most respondents (98.5%) reported having heard of NTDs, reflecting the reach of mass sensitisation campaigns. However, perceived causes of skin disease were predominantly related to hygiene (inadequate hygiene: 75.9%), diet and skin care (67.9%), and external environmental factors (61.2%). Notably, 22.8% attributed skin diseases to sociocultural factors such as witchcraft, divine punishment, or curses (Table 5).

**Table 5.**
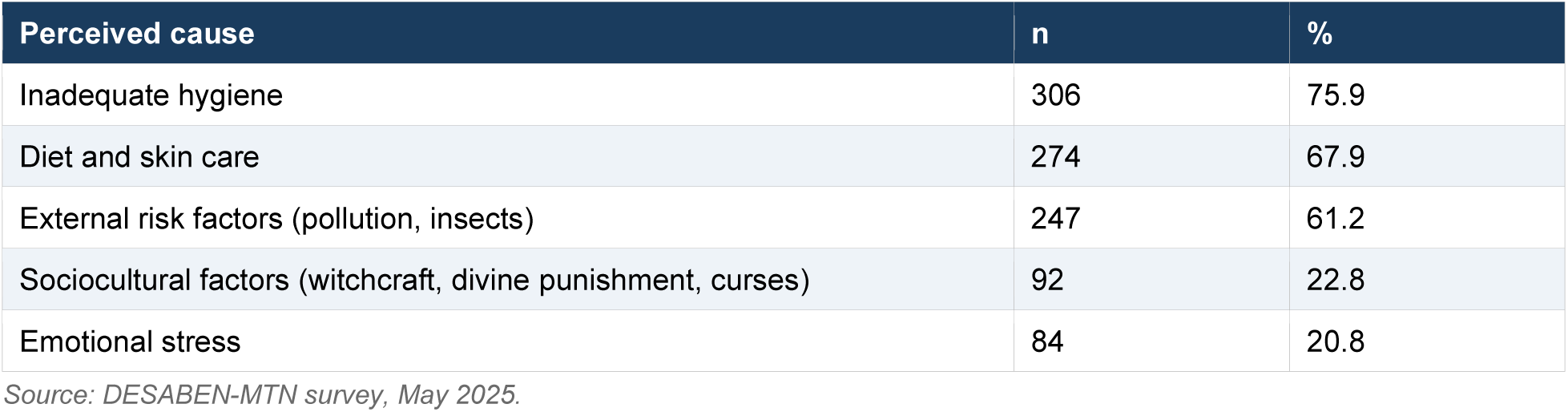
Perceived causes of skin diseases (multiple responses allowed, N=403).

Qualitative interviews confirmed that mystical interpretations of skin NTDs are widespread, particularly for slow-healing lesions. A relative of a patient in Zagnanado (36-40 years) stated: *“These chronic long-lasting wounds are not natural diseases. It is often an envouṫement (spell). You must consult the bɔkɔnɔ (traditional diviner) to understand what it is. You should not treat it in hospital, otherwise the person will die.”* These beliefs drive initial recourse to traditional healers and the use of plant-based preparations (*aizizɔn godonu, zomè, alomangbo, Houéflou*), substantially delaying biomedical consultation.

### Living conditions and environmental risk factors

Environmental factors were identified as major transmission determinants by 62.5% of participants (Allada: 64.1%; Zagnanado: 61.2%). Significant commune-level differences were noted for the predominant environmental risk type: precarious sanitation conditions dominated in Zagnanado (82.5% of respondents), whereas vector and insect exposure prevailed in Allada (60.9%), pointing to distinct epidemiological profiles that warrant differentiated responses.

The practice locally termed *“tout dans le même trou”* (all in the same hole) using a single water source for all domestic activities including bathing, cooking, laundering, and defecation was widely documented. Combined with inadequate wound care and lack of body hygiene after agricultural work, these practices sustain transmission. A health provider from Zagnanado (56-60 years) noted: *“Thatched-roof housing maintains humidity and fosters insect and bacterial proliferation. Most patients we receive live in unsanitised dwellings.”* Access to safe drinking water remains critically limited: in some localities, households travel up to 7 km to reach a functioning water point, forcing reliance on surface water and traditional wells.

### Proposed control strategies

Proposed strategies differed significantly between communes, reflecting their distinct needs (Table 6). Financial support for care access was the top priority in Allada (62.3% vs. 23.0% in Zagnanado; χ²=62.6, p<0.001, V=0.39), while improving health infrastructure (39.3% vs. 14.1%; χ²=33.5, p<0.001, V=0.29) and strengthening the health workforce (32.2% vs. 10.5%) were primary priorities in Zagnanado. Access to safe water and sanitation was similarly endorsed across communes (65.0% Allada vs. 57.4% Zagnanado; p=0.11). Integration of traditional medicine into control programmes received strong overall support (78.9%), with significantly higher endorsement in Allada (86.8% vs. 69.4%; χ²=18.2, p<0.001, V=0.21).

**Table 6.** Control strategies proposed by participants, by commune (multiple responses allowed). p-values from Pearson chi-squared tests. *p<0.001.

| Strategy | Allada n (%) | Zagnanado n (%) | Total n (%) | p-value |
| --- | --- | --- | --- | --- |
| Financial support for care access | 137 (62.3) | 42 (23.0) | 179 (44.4) | <0.001* |
| Improved health infrastructure | 31 (14.1) | 72 (39.3) | 108 (26.8) | <0.001* |
| Strengthened health workforce | 23 (10.5) | 59 (32.2) | 98 (24.3) | <0.001* |
| Safe water and sanitation | 143 (65.0) | 105 (57.4) | 248 (61.5) | 0.11 |
| Hygiene education | 68 (30.9) | 71 (38.8) | 139 (34.5) | 0.09 |
| Integration of traditional medicine | 191 (86.8) | 127 (69.4) | 318 (78.9) | <0.001* |

Source: DESABEN-MTN survey, May 2025.

## DISCUSSION

### A multidimensional vulnerability profile

Our study shows that people affected by skin NTDs in Zagnanado and Allada present a multidimensional vulnerability profile consistent with global NTD epidemiology. The predominance of farming activities (43.7%), low educational attainment (38.5% with no schooling), and high rural residence (50.1%) confirm WHO’s observation that NTDs primarily affect poor rural populations [12]. The informal nature of economic activities and instability of income markedly limit capacity to access care [13,14]. These findings resonate with profiles documented in Togo, Burkina Faso, and elsewhere in West Africa [12]. A Beninese specificity, however, is the dual exposure associated with aquatic livelihoods fishing, rice growing, and raffia collection which bring populations into sustained contact with hydromorphic environments where *Mycobacterium ulcerans* thrives.

### Nosological dissociation: a critical and under-recognised barrier

The finding that only 7.2% of participants spontaneously named Buruli ulcer despite both sites hosting specialised treatment centres is arguably the most important result of this study. We conceptualise this as ***nosological dissociation***: the perceptual separation between a disease’s biomedical identity and its local vernacular equivalents. Similar phenomena have been documented in Cameroon and Côte d’Ivoire, where patients arrive at CDTUBs at advanced ulcerative stages after exhausting traditional care pathways [18]. In the Beninese context, the Fongbé nomenclature assigns different names to different disease stages *akpa* for a generic wound, *akpa djɔmakou* for a chronic non-healing wound, *timantibo* for a verrucous lesion without connecting them to a single biomedical entity. This has direct programmatic implications: awareness campaigns that use only the term “Buruli ulcer” are unlikely to be effective. Effective communication must bridge vernacular and biomedical terminologies, for example by training community health workers to use local NTD names during outreach sessions and explicitly linking them to their biomedical equivalents. This approach has been piloted with modest success in Côte d’Ivoire’s Buruli ulcer programme and warrants systematic evaluation in Benin.

### Economic barriers: universal vs. structural

A key analytical contribution of this study is the distinction between universal financial barriers treatment cost (84.6%, no commune difference, p=0.43) and structural barriers that operate differentially: limited access (V=0.40, p<0.001) and inadequate infrastructure (V=0.47, p<0.001). This distinction has direct policy implications. Financial protection mechanisms (insurance, vouchers, conditional cash transfers) are needed in both communes. However, geographical access and service density are structural deficits concentrated in Zagnanado, requiring infrastructure investment and health workforce deployment rather than financial instruments alone. The crude OR of 10.15 for infrastructure inadequacy (Table 7) is particularly striking and suggests that the Zagnanado–Allada gap is not a matter of perception but of objectively different service landscapes. These findings parallel results from Ghana and Nigeria, where NTD-affected households spend 20–40% of annual income on care [15,16], and underscore the urgency of multi-pronged strategies combining financial and geographic accessibility.

**Table 7.**
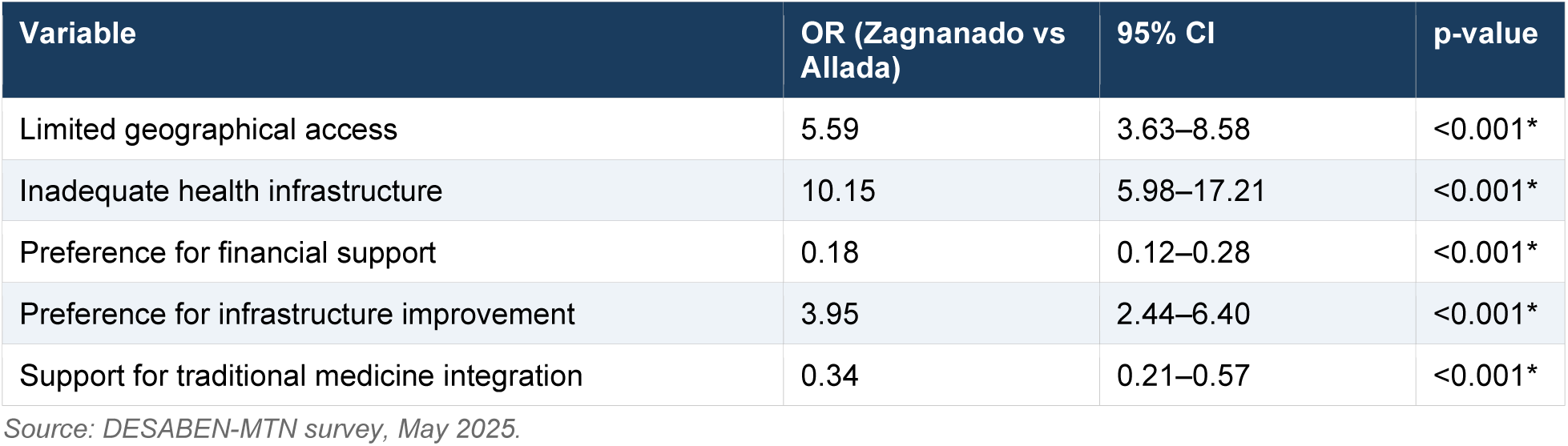
Crude odds ratios (OR) for access barriers and proposed strategies, Zagnanado vs. Allada (reference category). OR computed from 2×2 contingency tables (Woolf’s method). 95% CI = 95% confidence interval. *p<0.001 (Pearson chi-squared).

| Variable | OR (Zagnanado vs Allada) | 95% CI | p-value |
| --- | --- | --- | --- |
| Limited geographical access | 5.59 | 3.63–8.58 | <0.001* |
| Inadequate health infrastructure | 10.15 | 5.98–17.21 | <0.001* |
| Preference for financial support | 0.18 | 0.12–0.28 | <0.001* |
| Preference for infrastructure improvement | 3.95 | 2.44–6.40 | <0.001* |
| Support for traditional medicine integration | 0.34 | 0.21–0.57 | <0.001* |

Source: DESABEN-MTN survey, May 2025.

### Sociocultural barriers and traditional medicine

Mystical interpretations of skin NTDs (22.8% citing witchcraft or curses) constitute major barriers documented in other African contexts [17]. This quantitative finding was triangulated with and substantially enriched by qualitative data: the patient who refused hospital treatment for fear of death if treated biomedically (Zagnanado, 36-40 years, quoted in Results) illustrates a deeply ingrained belief system that no biomedical awareness campaign centred on germ theory can easily displace. The convergence of quantitative data (22.8% sociocultural attribution) and qualitative narratives suggests this represents a stable, socially shared illness representation rather than an idiosyncratic belief. The strong endorsement of traditional medicine integration (78.9% overall) should not be seen merely as a limitation to address, but as an asset to leverage. Given that traditional healers are the first port of call for most affected families, formalised collaboration frameworks as tested in leprosy and onchocerciasis programmes elsewhere could substantially shorten the delay between symptom onset and biomedical referral. A recent scoping review on community-based models for skin NTDs recommends integrating traditional healers to improve NTD awareness, reduce stigma, and enhance treatment adherence [20]. This is consistent with WHO’s 2019 traditional medicine strategy and recent inclusive NTD programme frameworks [21].

### One Health and occupational-environmental risk

The One Health approach embedded in our conceptual framework is particularly salient for explaining skin NTD persistence in these communes. Populations engaged in livestock rearing and fishing in hydromorphic zones are doubly exposed through direct contact with aquatic environments and through handling of animals or by-products under precarious hygiene conditions. Several participants reported handling livestock (cattle, goats, pigs) and fish in hydromorphic environments without protective equipment, compounding transmission risk at the human–animal–environment interface. Raffia collection in the swamps of Allada is an archetypical example that directly exposes women to *M. ulcerans* infection. The *tout dans le même trou* practice exemplifies a cross-contamination cycle sustained by inadequate water and sanitation infrastructure. Effective control programmes cannot be limited to the clinical dimension: they must integrate environmental management, food safety, and ecosystem protection. The One Health framework offers an integrative perspective for designing more comprehensive and ecologically adapted responses [10].

### Limitations

Several limitations must be acknowledged. First, the cross-sectional design precludes causal inference between socioeconomic determinants and skin NTD burden. Longitudinal cohort studies are needed to establish directionality and measure temporal changes in risk factors. Second, restriction to two communes may limit generalisability to other endemic areas of Benin with different geographic and socioeconomic profiles. Third, the mixed composition of the sample confirmed NTD cases and household relatives/community members introduces potential selection bias: CDTUB-registered patients may disproportionately represent more severe cases, while community recruits may include undetected cases. Future studies should explicitly stratify analyses by case vs. non-case status. Fourth, self-reported data are susceptible to recall bias, social desirability bias (particularly regarding health-seeking behaviour and income), and cultural bias (stigmatised conditions may be under-declared). Fifth, the quantitative analysis was limited to descriptive statistics and bivariate chi-squared tests; while full multivariate logistic regression was not feasible given the aggregate nature of data available, crude odds ratios from bivariate analyses are presented in Table 7 and provide directional evidence for the associations observed; multivariate analyses are planned for a multi-commune follow-up study.

## CONCLUSION

The DESABEN-MTN study provides novel, commune-level evidence on the socioeconomic and environmental determinants sustaining skin NTDs in Zagnanado and Allada, Benin. The affected population comprises predominantly rural adults with low educational attainment and informal livelihoods that expose them to NTD-conducive environments. Statistically significant intercommunal differences in care access barriers reflecting structural differences in health service density call for differentiated, context-specific strategies. The near-absence of Buruli ulcer recognition (7.2%), framed here as nosological dissociation, reveals a critical failure of current awareness campaigns that use biomedical terminology disconnected from vernacular illness concepts.

Breaking the disease–poverty cycle requires holistic interventions: financial protection mechanisms (universal in both communes); infrastructure strengthening (priority in Zagnanado); culturally adapted communication bridging Fongbé and biomedical disease names; structured collaboration with traditional healers; training of community actors; and sustained political investment in CDTUB capacity. These findings inform the WHO NTD Roadmap 2021–2030 and the PNLLUB’s national control strategy for skin NTDs in Benin.

## ACKNOWLEDGEMENTS

The authors thank the Fondation Anesvad and the Fondation Raoul Follereau for funding; all study participants; the local authorities of Zagnanado and Allada; the CDTUB staff; health professionals and traditional healers; and the full DESABEN-MTN research team.

## AUTHOR CONTRIBUTIONS

RH: Conceptualization, Funding acquisition, Methodology, Project administration, Supervision, Writing – review & editing (Lead). MB: Conceptualization, Data curation, Formal analysis, Investigation, Methodology, Software, Validation, Visualization, Writing – original draft, Writing – review & editing (Lead). BG: Data curation, Investigation, Validation, Writing – review & editing (Supporting). FT: Investigation, Data curation, Writing – review & editing (Supporting). DA: Investigation, Resources, Writing – review & editing (Supporting). All authors read and approved the final manuscript. Author roles follow the CRediT taxonomy (https://credit.niso.org/).

## DATA AVAILABILITY

The anonymised dataset and data collection instruments (quantitative questionnaire and qualitative interview guide, in French and English) are available at Zenodo: https://doi.org/10.5281/zenodo.19664736. Released under CC BY 4.0. Qualitative transcripts are available from the corresponding author on reasonable request, subject to approval from the National Ethics Committee for Health Research of Benin (CNERS).

## SUPPORTING INFORMATION

**S1 Checklist.** STROBE checklist for cross-sectional studies.

**S1 File.** Quantitative structured questionnaire (English version).

**S2 File.** Qualitative Semi-Structured Interview Guide (English version)

## REFERENCES

1. World Health Organization. Social determinants of health: key concepts [Internet]. Geneva: WHO; 2021 [cited 2025 May]. Available from: https://www.who.int/health-topics/social-determinants-of-health

2. World Bank Group. Poverty and shared prosperity 2022: correcting course. Washington, DC: World Bank; 2022. doi:10.1596/978-1-4648-1893-6

3. Hotez PJ, Molyneux DH, Fenwick A, Kumaresan J, Sachs SE, Sachs JD, et al. Control of neglected tropical diseases. N Engl J Med. 2007;357(10):1018–1027. doi:10.1056/NEJMra064142

4. World Health Organization. Neglected tropical diseases: situation report 2023. Geneva: WHO; 2023.

5. World Health Organization. Neglected tropical diseases: road map for 2021–2030. Ending the neglect to attain the sustainable development goals. Geneva: WHO; 2021.

6. Programme National de Lutte contre la Lèpre, l’Ulcère de Buruli et le Pian (PNLLUB). Rapport de monitoring de l’Ulcère de Buruli 2023. Cotonou: Ministère de la Santé du Bénin; 2023.

7. Houndjrèbo FA. Les ulcères chroniques au Sud du Bénin : logiques et pratiques de prise en charge d’une maladie tropicale négligée à manifestation cutanée [doctoral thesis]. Abomey-Calavi: Université d’Abomey-Calavi; 2019. 300 p.

8. Institut National de la Statistique et de l’Analyse Économique (INSAE). Quatrième Recensement Général de la Population et de l’Habitation (RGPH4). Cotonou: INSAE; 2013.

9. Solar O, Irwin A. A conceptual framework for action on the social determinants of health. Social Determinants of Health Discussion Paper 2 (Policy and Practice). Geneva: World Health Organization; 2010. 79 p.

10. World Organisation for Animal Health (WOAH/OIE). One Health: a new professional imperative [Internet]. Paris: WOAH; 2022 [cited 2025 Jan]. Available from: https://www.woah.org/en/what-we-do/global-initiatives/one-health/

11. Creswell JW, Plano Clark VL. Designing and conducting mixed methods research. 3rd ed. Thousand Oaks, CA: SAGE Publications; 2018. 520 p.

12. World Health Organization. Neglected tropical diseases: progress report 2000–2022 and road map targets for 2030. Geneva: WHO; 2022.

13. Farmer PE. Pathologies of power: health, human rights, and the new war on the poor. Berkeley, CA: University of California Press; 2003. 402 p.

14. Mikkonen J, Raphael D. Social determinants of health: the Canadian facts. Toronto: York University School of Health Policy and Management; 2010. 62 p.

15. Aheto JMK, Amoah B, Koram KA, Fobil JN, Gordon A. Predictors of access and utilisation of health services and financial risk among people affected by NTDs in Ghana. PLoS Negl Trop Dis. 2020;14(7):e0008489. doi:10.1371/journal.pntd.0008489

16. Ekpo UF, Oluwole AS, Ojo OO, Oyibo WA, Fajana OF, Mafiana CF. Soil-transmitted helminths and financial burden of care among school-age children in Nigeria. Acta Trop. 2012;121(3):193–199. doi:10.1016/j.actatropica.2011.12.004

17. Toomer K, Nkesa L, Winkler A, Boateng GO. Stigma and the social determinants of health in neglected tropical diseases: a scoping review. Soc Sci Med. 2019;241:112527. doi:10.1016/j.socscimed.2019.112527

18. Tchatchouang S, Piubello A, Gody JC, Eyangoh S, Marsollier L, Marion E. Integrated active case detection and management of skin NTDs in yaws-endemic health districts: a cross-sectional study. PLoS Negl Trop Dis. 2024;18(2):e0011980. doi:10.1371/journal.pntd.0011980

19. World Health Organization. Report of the WHO advisory group meeting on Buruli ulcer. Geneva: WHO; 2019. WHO/CDS/NTD/IDM/2019.01.

20. Ngangue P, Bedard M, Lussier MT, Karemere H, Louchard A, Ganete TK. Community-based approaches for skin NTD control: a scoping review of methods and outcomes. PLoS Negl Trop Dis. 2015;9(12):e0004174. doi:10.1371/journal.pntd.0004174

21. World Health Organization. WHO global strategy for traditional and complementary medicine 2019–2025. Geneva: WHO; 2019. Licence: CC BY-NC-SA 3.0 IGO.

